# A review of reviews exploring patient and public involvement in population health research

**DOI:** 10.1101/2022.11.16.22282319

**Authors:** Soo Vinnicombe, Jane Noyes

## Abstract

**Introduction:** Patient and public involvement (PPI) is increasingly seen as something that is integral to research and of importance to research funders. There is general recognition that PPI is the right thing to do for both moral and practical reasons. The aim of this review of reviews is to examine how PPI can be done ‘properly’ by looking at the evidence that exists from published reviews and assessing it against the UK Standards for Public Involvement in Research, as well as examining the specific features of population health research that can make PPI more challenging.

**Methods:** A review of reviews was carried out following the 5-stage Framework Synthesis method.

**Results:** In total 31 reviews were included. There is a lack of current research or clarity around Governance and Impact when findings are mapped against UK Standards for Public Involvement in Research. It was also clear that there is little knowledge around PPI with under-represented groups. There are gaps in knowledge about how to ensure key specific attributes of population health research are addressed for PPI team members – particularly around how to deal with complexity and the data-driven nature of the research. Two tools were produced for researchers and PPI members to further improve their PPI activity within population health research and health research more generally: A framework of recommended actions to address PPI in population health research, and guidance on integrating PPI based on the UK Standards for Public Involvement in Research.

**Conclusions:** Facilitating PPI in population health research is challenging due to the nature of this type of research and there is far less evidence on how to do PPI well in this context. The tools can help researchers identify key aspects of PPI that can be integrated when designing PPI within projects. Findings also highlight specific areas where more research or discussion is needed.

## Introduction

The focus of this review of reviews is on Public and Patient involvement (PPI) in population health research. PPI is increasingly seen as something that is integral to research and of importance to research funders. For our purposes, PPI is defined as:

‘Public involvement in research means research that is done ‘with’ or ‘by’ the public, not ‘to’, ‘about’ or ‘for’ them. It means that patients or other people with relevant experience contribute to how research is designed, conducted and disseminated. It does not refer to research participants taking part in a study. Public involvement is also different from public engagement, which is when information and knowledge about research is shared with the public (1). There is however sometimes confusion between what constitutes public engagement compared with involvement. In some countries, such as Canada, it is also common to use ‘public engagement’ to refer to public involvement (2). Similarly, the lines between stakeholder representation and public or patient representation can sometimes be blurred.

### Population health research

‘Population health’ is associated with several definitions and nuances and there is overlap with public health and aspects of more general health research. The King’s Fund describes population health as:

‘An approach aimed at improving the health of an entire population. It is about improving the physical and mental health outcomes and wellbeing of people within and across a defined local, regional or national population, while reducing health inequalities. It includes action to reduce the occurrence of ill health, action to deliver appropriate health and care services and action on the wider determinants of health. It requires working with communities and partner agencies (3).

Public health, by comparison, can be defined as:

‘Activities to strengthen public health capacities and service aim to provide conditions under which people can maintain to be healthy, improve their health and wellbeing, or prevent the deterioration of their health. Public health focuses on the entire spectrum of health and wellbeing, not only the eradication of particular diseases (4).

Some refer to Public Health (note the capitalisation) as specifically about activities and interventions carried out by government agencies, health professionals, or other centralised bodies whereas population health includes other, non-health related, influences such as housing, transport and education. In reality, these various definitions can oversimplify our understanding and a rigid adherence to a perceived difference between the terms may serve to disguise relevant information about successful PPI activity. As Diez-Roux argues, ‘Much of this is a semantic discussion. What really matters are the questions we pose regarding the health of the public, the answers we obtain, and the actions we take in response. Whether we call this approach public health or population health is, in all honesty, irrelevant (5).

### Specific challenges of integrating PPI in population health research

Population health research, or health research that considers population level questions, provides challenges in terms of PPI that are not always present in condition-specific research projects. For example:

- Duration. Population health research often looks at health variables across a long period of time. This makes recruiting and retaining suitable PPI representation across the length of the project more challenging. Changes in personnel, in all parts of the research team and partners, can be expected in any project.
- Complexity. Population health is often multi-disciplinary and looks at health as the product of multiple determinants (such as biology, genetics, behaviours, social and environmental aspects) as well as looking at their interactions among individuals and groups and across time and generations. With all these different variants involved it can be difficult for a lay person to understand the complexity – or, to put it another way, for the researchers to explain the research in a way that a lay person can understand. It may often be the case that a different skill set, and therefore potentially a different person, is necessary at different stages of the research or for different workstreams – something that applies to researchers as well as to PPI representatives.
- Data-driven. Population health projects are often driven by large datasets and can involve knowledge of algorithms, advanced statistics, and analytical techniques that can be unfriendly to the non-mathematically minded. It can be a challenge for researchers to ‘translate’ both the process and the outcomes of their research in terms that can be more widely understood. This is one reason why PPI can be so helpful in such projects. For example, helping to design dissemination activity that is meaningful to a broad audience.
- Representation. Population health research often addresses large and diverse population groups within the populations being researched, which raises issues about the PPI being representative. Even within disease-specific studies it is often difficult, if not practically impossible, to recruit someone who truly represents the breadth of people with a certain condition. Once that issue is expanded out to wider populations, the issue of true representation is multiplied many times. Representation becomes particularly difficult with certain demographic groups which may be grouped together for convenience, but which might hide a variety of differences. A prime example of this is the involvement of Black, Asian and Minority Ethnic (BAME) individuals – recruiting a single person of BAME background risks subsuming important differences according to specific cultural, genetic, class, education and other factors. There is also an ongoing debate about terminology such as ‘hard to reach’, ‘under-represented’, ‘seldom heard’ and ‘under-served’ which often have problematic resonances (6). One way to think of this is that ‘[t]he key idea here is that the definition of ‘under-served’ is highly context-specific; it will depend on the population, the condition under study, the question being asked by research teams, and the intervention being tested. No single, simple definition can encompass all under-served groups (7).

### The need for a review of reviews

As described above, population health presents specific challenges for researchers and there is a lack of guidance on doing PPI well in population health research. Scoping searches identified a number of reviews of PPI involvement covering population health, public health as well as other more general reviews that included population and public health studies of interest. None of the published reviews had a specific focus on what worked to deliver optimal PPI in population health research. We therefore decided to undertake a review of reviews to explore the challenges and solutions to carrying out PPI well in population health research. Findings were then developed into two tools for researchers and PPI members to further improve the quality of PPI activities in population health research.

## Materials and methods

This review of reviews assembled and interpreted the evidence on PPI involvement in population health research. Question formulation was underpinned by the ECLIPSE (**E**xpectation, **C**lient Group, **L**ocation, **P**rofessionals and **Se**rvice) framework that is acknowledged to be most suitable for searching for health policy or health management information (8).

We developed the following question: What evidence exists concerning the successful development, implementation and evaluation of patient and public involvement activity or models in population health research in the UK and equivalent health systems?

### Inclusion criteria

- Type of study: systematic and other reviews that focus on the concept of, or approaches to, PPI and/or PPE (patient and public engagement) across population health, public health, health and social care. Limited to systematic reviews, narrative reviews, literature reviews, bibliometric reviews, scoping reviews and meta-analyses. Quantitative, qualitative and mixed-methods reviews were of interest.
- Setting: any organisational setting that includes population health, public health, health or social care aspects (e.g., primary care, mental health, hospital, tertiary care, voluntary, etc.).
- Type of involvement: not just being part of the research as a participant but being involved in part or all of the following stages – research development, research monitoring, research analysis and dissemination.

### Exclusion Criteria

- Articles not in English.
- Reviews published before 2010. However, the timeframes for the primary studies included in the reviews vary and can go back to the inception of various databases. This timeframe was considered appropriate as public and patient involvement is something that has been developing rapidly in recent years and was not really established as a well-recognised term before then.

### Search Strategy

An information scientist undertook the initial search of the Medline and PubMed databases. The full search strategy is included in Supporting Information 1. The Involve Evidence Library was searched for ‘systematic reviews’. Note that this library currently (September 2021) only includes references up to 2015.

The original search was done in May 2020 with a follow up search (stages 2 and 3) carried out early in September 2021 to pick up new reviews up to end of August 2021.

### Screening

Titles and abstracts were screened to identify reviews that met the inclusion criteria. Potentially relevant reviews were retrieved and the full text assessed for inclusion (Figure 1). The process was undertaken by SV and independently checked by JN.

**Figure 1.**
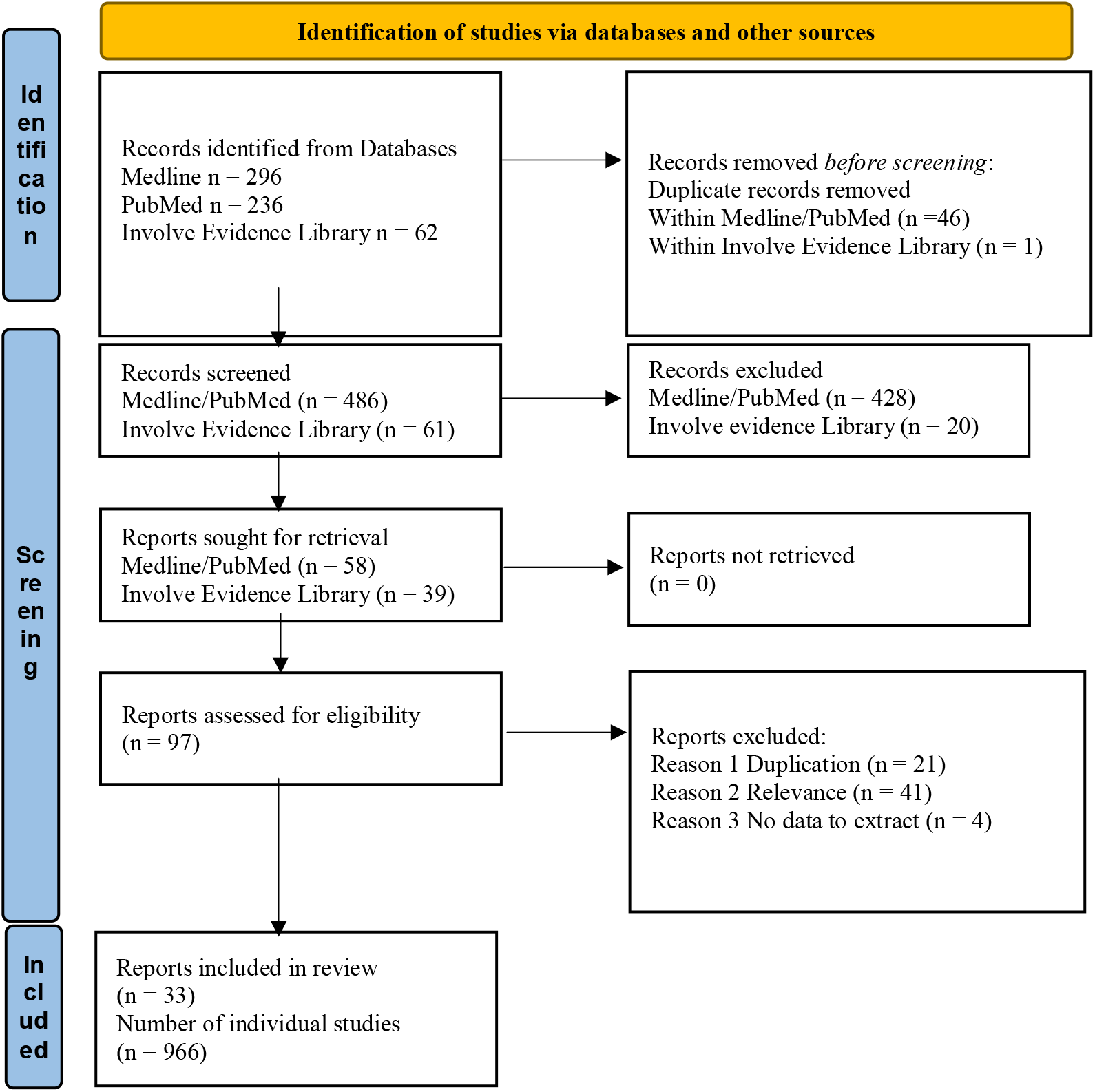
PRISMA Flow Diagram * This number is incomplete due to missing information on some papers. Duplicates have been removed original n = 1222).

### Quality appraisal

Originally the AMSTAR2 (9), method was trialled on six reviews but as most of the included reviews were qualitative rather than quantitative many of the AMSTAR2 domains did not apply so we switched to using CASP for systematic reviews (10). Included reviews were quality appraised by SV and independently checked by JN (Supporting Information 2). Reviews were not excluded at this stage on methodological grounds as the focus was on PPI processes reported in the review.

### Data extraction and synthesis

Studies included in source reviews were mapped for duplication and this was taken account of in the analysis and synthesis. As this review of reviews did not require a transformative method of data synthesis to better understand the descriptive accounts of PPI in the source reviews, we selected the aggregative 5-stage framework synthesis method for integrating evidence of interest from diverse review designs.

It is a matrix-based method involving the construction of a priori thematic categories into which data can be coded. The five stages are:

- Familiarisation
- Identifying a thematic framework
- Indexing
- Charting
- Mapping and interpretation (11)

Initial data extraction was carried out against a framework designed by the authors based on close examination of background literature and initial review readings (Table 1).

**Table 1.** Initial framework: headings and details.

| Main info | Title<br>Authors |
| --- | --- |
| Extracted information | Year published<br>Type of review<br>Area of focus<br>No. of studies<br>No. of papers<br>Full list<br>Databases searched<br>Other searches<br>Years searched<br>Exclusions<br>Geography<br>Methods used<br>Included PPI in own review |
| Why do PPI? | Attribute<br>Who benefits?<br>Evidence for<br>Evidence against |
| How to do PPI – especially in population health research | Attribute – barrier<br>Stage affected<br>Mitigation<br>Attribute - facilitator<br>Stage affected<br>Good practice |
| Terminology | Types of PPI<br>Stages of research<br>Other |
| Other | Gaps in Knowledge<br>Country specific legislation/ guidance<br>Case studies? |

Extracted data were subsequently mapped against a second framework (Table 2) and matched against the UK Standards for Public Involvement.

**Table 2.** Secondary framework: thematic mapping.

| Challenges |  |  | Solutions |  |  |
| --- | --- | --- | --- | --- | --- |
| Study id | Problem | Consequence | Study id | Solution | Details |

The UK Standards for Public Involvement are:

- Inclusive Opportunities - Offer public involvement opportunities that are accessible and that reach people and groups according to research needs.
- Working Together - Work together in a way that values all contributions, and that builds and sustains mutually respectful and productive relationships.
- Support and Learning - We offer and promote support and learning that builds confidence and skills for public involvement in research.
- Governance - Involve the public in research management, regulation, leadership and decision making.
- Communications - Use plain language for well-timed and relevant communications, as part of involvement plans and activities.
- Impact - Seek improvement by identifying and sharing the difference that public involvement makes to research (12).

### Public and patient involvement

This review of reviews was discussed with the Centre for Population Health Patient and Public Involvement Advisory Group, which meets quarterly to help set the strategic direction for PPI within the Centre. The draft review was read and commented on several times throughout its development by two PPI advisory group members.

## Results

Thirty-one reviews were included covering around one thousand individual studies, which were mainly based in the UK or USA. We took note of any duplication of studies across reviews to ensure that we were not double counting the evidence.

The studies covered a range of settings and subject areas (Supporting Information 3). Reviews varied in quality (Supporting Information 2) but as the review methods and findings were not the primary phenomenon of interest we did not place a lot of emphasis on the quality of the source reviews when interpreting findings.

Specifically, the reviews covered, to varying degrees, three out of the four challenges, outlined earlier, that set population health research apart from many other research types.

**Representation** was extensively discussed in the studies reviewed. It is an aspect of PPI that does not have a simple solution for any type of research project. For population health projects that tend to be longer in duration, it may be that different people need to take part in different periods of the project and, for complex projects, that different people need to be involved in different work streams. Boote (13) notes that there is a concern that PPI representatives taking part in research over time may become ‘professionalised’ and come to see things from the point of view of the research team rather than as a member of the public or patient demographic.

**Complexity** was also discussed when talking about support and learning requirements for PPI members. Population health projects are often highly complex but, given the right support and training, that is not a sufficient reason to exclude PPI activity.

The **data-driven** aspect was touched upon mainly in terms of ensuring that project specific training and support was available. Many population health projects include aspects of Big Data which can add a layer of difficulty to PPI activity, but which can also be addressed by considering tailored training and support. Having non-data experts involved in such projects may help when designing dissemination and communication activities around the project so that they can eventually be more accessible to a wider audience.

**Duration** was the only aspect that was not specifically discussed in the reviews and in finding solutions. It is possible to postulate that building relationships and strong ways of working together may help to address this issue. But also, that acknowledging upfront the changing requirements of a long-term project will help researchers to plan accordingly – including planning for long term PPI.

### Common issues across PPI activity in population and other types of health research

There are several aspects of PPI activity that are common across various types of health research, including, but not exclusive to, population health research.

### Challenges

Just over half of the reviews (18 out of 31 (14-31)) noted a range of potential challenges with PPI that were reported to stand in the way of the successful development, implementation and evaluation of patient and public involvement activity or models in health research in the UK and equivalent health systems.

Consolidation of the challenges reported in the reviews suggested that the following (Table 3) were the key issues. These have been grouped into appropriate headings.

**Table 3.** Full list of challenges identified.

| Heading | Sub-heading | Reviews |
| --- | --- | --- |
| **Resources** | Lack of budget | 14, 16, 18, 19, 20, 25, 27, 28 |
|  | Lack of time | 14, 16, 18, 19, 20, 25, 27 |
|  | Emotional burden on PPI members | 14, 20, 21, 25 |
|  | Complicated logistics/ infrastructure | 16, 19 |
|  | Workload too high (on all sides) | 20, 23 |
|  | Lack of incentives | 16 |
|  | Lack of preparation | 14 |
|  | Lack of staff continuity | 16 |
|  | Lack of support for PPI members | 24 |
|  | Scope creep<sup>1</sup> | 26 |
| **Conflict and control** | Allowing power to be shared with PPI | 14, 15, 16, 17, 19, 21, 22, 25 |
|  | Expectations (from all sides) | 14, 16, 20, 21, 27, 29 |
|  | Conflicting perspectives | 15, 16, 19, 23, 24 |
|  | A culture of researchers vs PPI members | 14, 16, 20 |
|  | Ethical concerns | 24, 25 |
|  | Challenging the establishment | 14 |
|  | Differences within communities | 14 |
|  | Accepting the legitimacy of PPI | 19 |
|  | Prioritising personal experience | 18 |
|  | Scepticism (from all sides) | 14 |
|  | Unresolved conflict | 31 |
| **Knowledge** | Processes | 14, 16, 19, 25, 27 |
|  | Language/ jargon | 14, 15, 18, 19, 27 |
|  | Lack of skills or training | 14, 19, 23, 24, 25 |
|  | Administration issues | 17 |
|  | Working practices | 14 |
| **Representation** | Reflecting the diversity of affected populations | 14, 17, 18, 19, 23, 25, 27, 30 |
|  | Tokenism of PPI (aka box-ticking) | 22, 24, 26 |
|  | Getting early-stage involvement | 17, 22 |
|  | Involving children | 19 |
|  | Protecting anonymity | 25 |
|  | Accessibility (venues) | 28 |
| **Communication** | Lack of meaningful and timely communication leading to disenfranchisement | 14, 17, 21 |
|  | Difficulty reporting impact of PPI | 15, 24, 25 |
|  | Building relationships to sustain involvement | 16, 19 |
|  | Transparency of research process | 23 |
|  | Building trust (on all sides) | 16 |
|  | Different values within team | 27 |

Many of these challenges will be even more apparent in population health research where projects tend to face the four challenges of: longer duration, involving more complex and varied processes, alongside issues of big data, and finding appropriate representation to cover the project breadth and length.

### Solutions

Nearly three quarters of the studies (23 out of 31) (2, 16-18, 20-38) noted a range of potential solutions for ensuring that PPI was more likely to be successful.

These proposed solutions have been collated, consolidated and sorted according to the UK Standards for Involvement in Research as follows:

### Inclusive Opportunities

#### Solution

Offer public involvement opportunities that are accessible and that reach people and groups according to research needs. Research also needs to be informed by a diversity of public experience and insight, so that it leads to treatments and services which reflect these needs.

Eleven reviews mentioned inclusion (17-18, 20-21, 24, 29, 33-34, 36-38). Key themes emerged from these studies and these are outlined in Table 4 below and explicitly addressed the problem area of Representation.

**Table 4.**
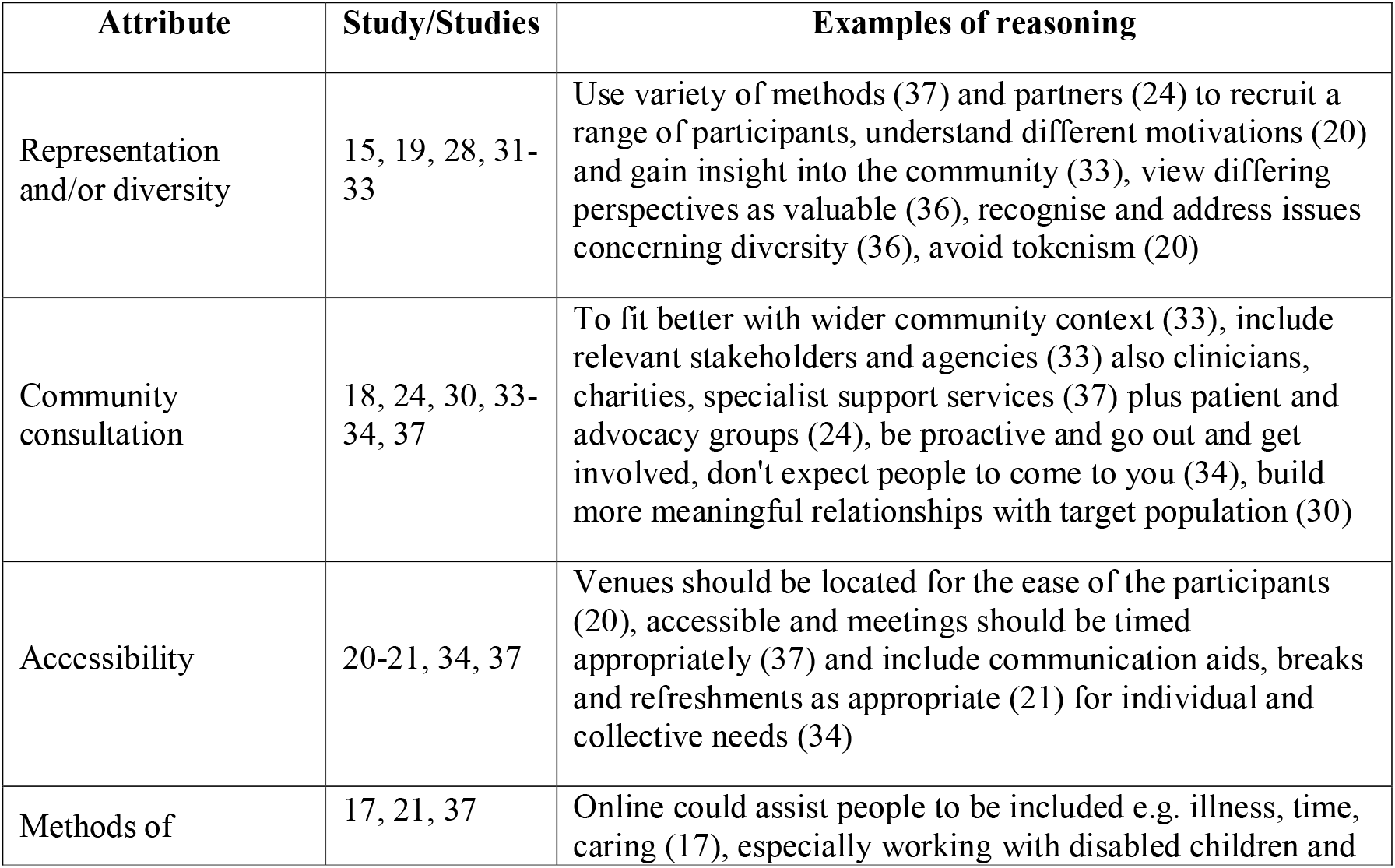
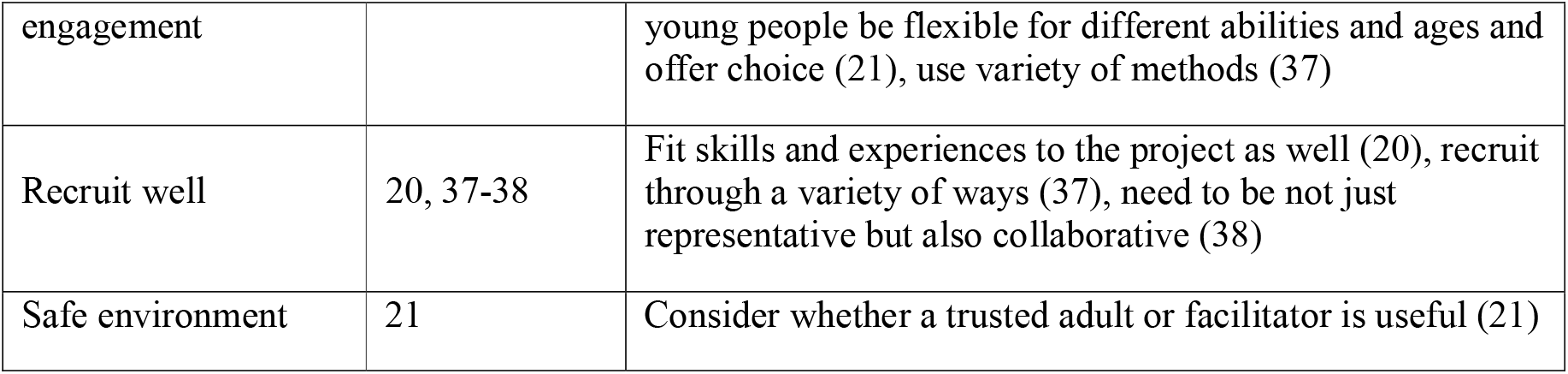
Solutions – Inclusive opportunities.

### Working Together

#### Solution

Work together in a way that values all contributions, and that builds and sustains mutually respectful and productive relationships. Public involvement in research is better when people work together towards a common purpose, and different perspectives are respected.

Twenty-one reviews (2, 16-18, 20-21, 23-29, 31-38) discussed aspects of this standard. The main areas of discussion are outlined in Table 5 below and explicitly addresses the problem area of Conflict and Control.

**Table 5.**
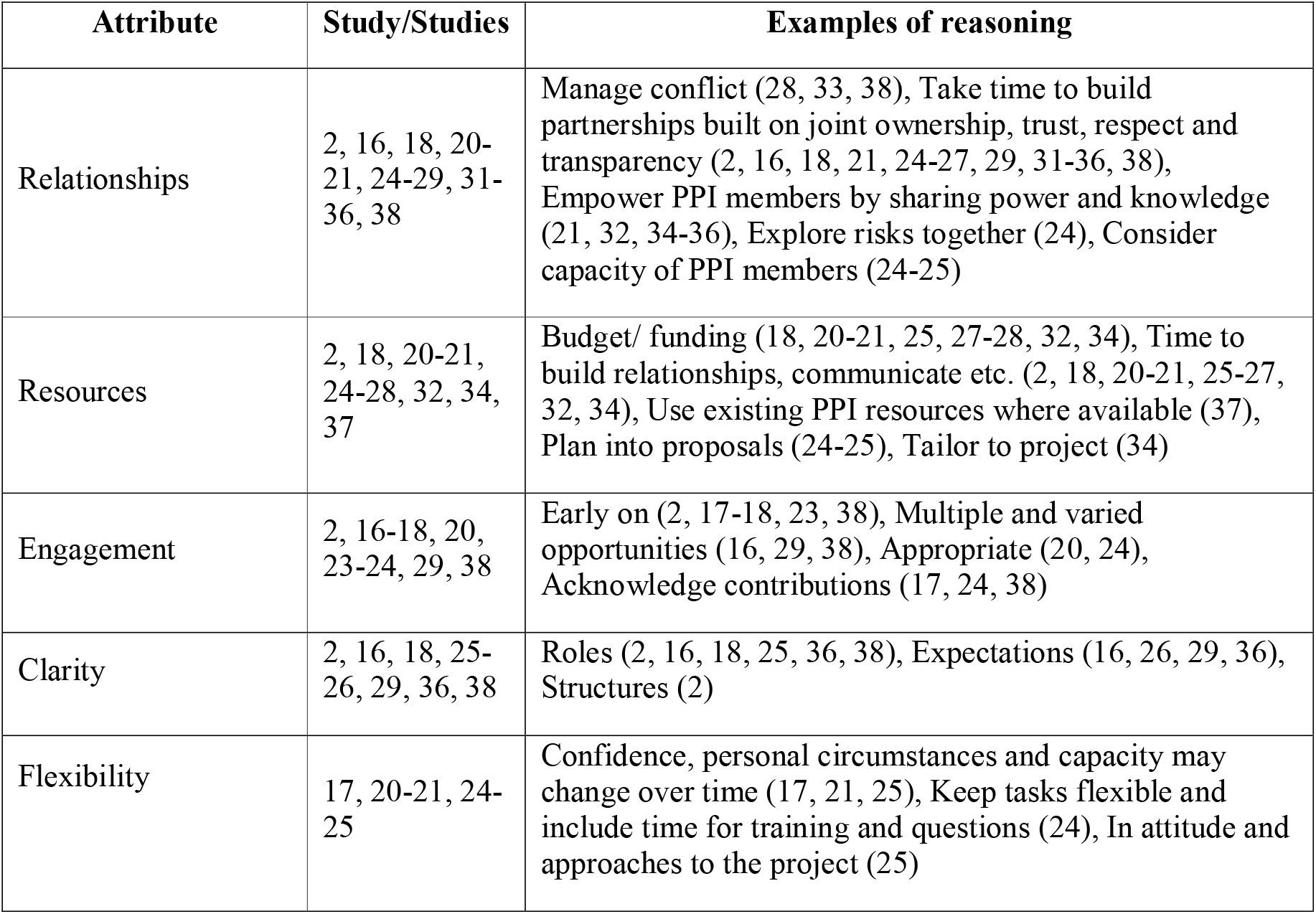
Solutions – Working Together.

| Attribute | Study/Studies | Examples of reasoning |
| --- | --- | --- |
| Relationships | 2, 16, 18, 20-21, 24-29, 31-36, 38 | Manage conflict (28, 33, 38), Take time to build partnerships built on joint ownership, trust, respect and transparency (2, 16, 18, 21, 24-27, 29, 31-36, 38), Empower PPI members by sharing power and knowledge (21, 32, 34-36), Explore risks together (24), Consider capacity of PPI members (24-25) |
| Resources | 2, 18, 20-21, 24-28, 32, 34, 37 | Budget/ funding (18, 20-21, 25, 27-28, 32, 34), Time to build relationships, communicate etc. (2, 18, 20-21, 25-27, 32, 34), Use existing PPI resources where available (37), Plan into proposals (24-25), Tailor to project (34) |
| Engagement | 2, 16-18, 20, 23-24, 29, 38 | Early on (2, 17-18, 23, 38), Multiple and varied opportunities (16, 29, 38), Appropriate (20, 24), Acknowledge contributions (17, 24, 38) |
| Clarity | 2, 16, 18, 25-26, 29, 36, 38 | Roles (2, 16, 18, 25, 36, 38), Expectations (16, 26, 29, 36), Structures (2) |
| Flexibility | 17, 20-21, 24-25 | Confidence, personal circumstances and capacity may change over time (17, 21, 25), Keep tasks flexible and include time for training and questions (24), In attitude and approaches to the project (25) |

### Support and Learning

#### Solution

Offer and promote support and learning that builds confidence and skills for public involvement in research. Seek to remove practical and social barriers that stop members of the public and research professionals from making the most of public involvement in research.

The following seventeen reviews mentioned various aspects of support and learning (2, 16, 18, 21-22, 24-25, 27-29, 32-38). The results are shown in Table 6 below. The table is split into two sections to reflect differences between support and learning methods, and explicitly addresses the problem area of Knowledge.

**Table 6.** Solutions – Support and learning.

| **SUPPORT - Attribute** | **Study/Studies** | **Examples of reasoning** |
| --- | --- | --- |
| Emotional support | 2, 13, 19, 24, 28-29, 31-32 | Recognise that experiences may be upsetting (18), Provide safe spaces (33), Provide consistent feedback and support (24), Consider how to deal with anxiety (29) |
| Practical support | 24, 34-36 | Think about details e.g. childcare, food, location, transport, compensation, timings (35), Have strategies for when people are ill/ can't take part (24) |
| Structural support | 16, 25, 36 | Make sure key project individuals support PPI (16), Provide structures that support PPI (36), Include relevant institutions such as charities, volunteer groups etc. (25) |
| Specific support | 29, 33 | Ensure support specific to topic area (29) and to their individual involvement (33). |
| **LEARNING - Attribute** | **Study/Studies** | **Examples of reasoning** |
| As appropriate | 2, 18, 27, 32-33, 36-38 | Make learning relevant to the specific context of the research (2, 27, 33) and at the appropriate level for the PPI member (33) to allow full participation (38) and to build participant capacity (18) |
| Formal knowledge | 16, 25, 32, 34 | Formal development of knowledge and skills (16), supporting participants to be informed and make informed decisions (25) and to understand specific parts of the research process and/or context (32) |
| Research methods | 22, 32, 37-38 | Training in research components to give confidence in their involvement (32) and to explain 'rules' and constraints of |
|  |  | research (22) |
| Variety of learning methods | 24, 29, 34-35 | Use a variety of methods such as supervision, mentoring, formal, workshops and team based (35), include everyone on the team if possible (24, 34) |
| Share knowledge | 32-33 | Acknowledge that knowledge and experience flow both ways and make ways to facilitate that flow (33) |
| General | 21, 25, 28, 34 | Provide, support and fund training and learning opportunities (25). |

### Governance

#### Solution

Involve the public in research management, regulation, leadership and decision making.

Public involvement in research governance can help research be more transparent and gain public trust. This section explicitly addresses the problem area of Conflict and Control. Only three of the reviews mentioned governance (2, 24, 35). They discuss the need for shared decision making (at every level), power and leadership, in order to lead to a culture of deeper involvement. As limited suggestions were reported there is no table for this section.

### Communications

#### Solution

Use plain language for well-timed and relevant communications, as part of involvement plans and activities. Communicate with a wider audience about public involvement and research, using a broad range of approaches that are accessible and appealing.

Nine of the reviews discussed communication as being important to ensure PPI activity is successful (2, 20, 24-25, 27, 32, 34-35, 37). Various attributes of good communication are discussed with the main points as in Table 7 below, and explicitly addresses the problem area of Communications.

**Table 7.** Solutions - Communications

| Attribute | Study/Studies | Examples of reasoning |
| --- | --- | --- |
| Listen, act and feed back | 24, 27, 34-35 | Helps address issues such as power (35), let people know what you are doing with their suggestions and why (24), ensures accountability (27) |
| Ongoing/ regular updates | 25, 32, 37 | Contribute to motivation and engagement, and |
|  |  | to foster satisfying partnerships (32) |
| Creating space to voice concern/ open communication climate | 20, 24, 32 | Contribute to motivation and engagement, and to foster satisfying partnerships (32) |
| Avoid/ translate jargon | 24-25, 32 | Ensuring everyone understood and felt comfortable and confident to engage in meaningful dialogue (32) |
| Use different materials (not just written reports etc) | 32, 34, 37 | Ensure people with different levels of literacy can participate (32) |
| Sharing information, experiences and knowledge | 2, 34 | Across all groups involved (2) |
| Clarifying and agree expectations upfront | 24, 32 | Could avoid conflicts, demotivation, dissolution of partnerships, or frustration in situations where stakeholders could perceive a lack of concrete actions (32), patients are 'partners' not 'are involved' (24) |
| Have stakeholders lead groups | 32 | But be careful they include all groups in the discussion (32) |

### Impact

#### Solution

Seek improvement by identifying and sharing the difference that public involvement makes to research. Understand the changes, benefits and learning gained from the insights and experiences of patients, carers and the public.

Seven of the reviews discussed impact (2, 20, 24, 32, 34-35, 38). The general theme was that impact needs to be better evaluated throughout the whole research lifecycle. It was noted that this is an area where the existing literature is scant and current working practices are perceived to be lacking in terms of rigour.

Evaluating impact through continuous assessment and feedback was seen to be important in order to ensure ongoing involvement, to identify best practice and areas for improvement, and to make sure that the experience is working for everyone involved.

In addition to evaluating the process of PPI within health research, it was also noted that the impact of findings that are translated to real world setting, and ideally the contribution of PPI activity to that impact, should also be evaluated.

It is important to note that impact can be positive or negative and that impact may happen in a complex way and to a range of areas, for example, impact on the research, on the research outcomes, on the researchers, on the PPI members, on the wider community and stakeholders.

### Other issues

Interestingly considering the topic of the reviews, the use of PPI members in the reviews was not universal.

- 9 reviews described PPI throughout the review process
- 3 reviews took their findings to PPI members for discussion
- 3 reviews made use of external panels or organisations
- Single reviews reported utilising PPI at specific stages:
  - To identify research questions
  - Reviewing protocol
  - During execution and translation
  - Reviewing the process
  - Feedback from stakeholder but stage not stated
- 2 reviews mentioned that there had not been any PPI in the review
- 9 reviews did not mention PPI in their own review process at all.

Few of the reviews detailed the studies discussed within them in terms of types of PPI or in terms of stages of research although most included some discussion of these areas in general terms. Dawson et al (39) is one exception where the studies are clearly detailed in terms of what PPI groups or individuals were involved in various tasks.

There was no consistent terminology used for either types of PPI or stages of research. There has been some attempt to categorise these at a national level. For example, in the UK, INVOLVE distinguished between three PPI approaches: consultation, collaboration and user-led; while Health Canada divides PPI into five stages: inform or educate, gather information, discuss, engage and partner (Pii)(40).

Crocker et al (41) describes the types of involvement covered in the studies to range ‘from one person to many people or whole patient organisations, from one-off involvement in a particular aspect of the trial (for example, reviewing draft information for patients or recruiting participants from their communities) to involvement throughout the trial (for example, as members of a trial steering committee), and from involvement with no decision making power (for example, as advisers) to involvement in decision making as equal partners’.

Some examples of stages of research where PPI could be included were described as below in Table 8.

**Table 8.** Examples of stages of research.

| Wilsher (23) | Domecq (26) | Pii (40) |
| --- | --- | --- |
| Identify/prioritise<br>Design<br>Grant development<br>Undertake/Manage<br>Analysing/interpret<br>Dissemination<br>Monitoring/evaluation | 1) Preparatory phase (agenda setting, prioritization of research topics and funding).<br>2) Execution phase (study design & procedures, study recruitment, data collection, and data analysis).<br>3) Translation phase (dissemination, implementation, and evaluation). | 1. Development of research focus<br>Research definition<br>Research prioritization<br>2. Development of research design<br>Method development<br>Study design development<br>3. Recruitment<br>Recruitment strategy<br>Recruitment<br>4. Data generation<br>5. Data processing/ Analysis<br>6. Research dissemination<br>Dissemination<br>Dissemination strategy |

## Discussion

### Population Health

This review of reviews set out to see what evidence there was concerning optimising patient and public involvement specific to population health research. The novelty in this review of reviews is twofold: firstly, that the findings have been framed by the UK Standards and secondly, that the challenges have been matched against potential solutions. Most reviews were about PPI activity in specific thematic healthcare areas or in general health and social care research but the details of the studies included in the reviews makes it clear that many studies included were of direct relevance to population health research. The findings are, therefore, both generic across health and social care research as well as providing useful evidence-based suggestions as to what works in PPI in population health research.

### Comparing findings with recently published primary studies

Looking at recently published primary studies we found several of interest, mainly around data-driven population health research. The principles that emerge from these studies fit well with the findings of the review of reviews, but also suggest that there are a variety of approaches through which PPI can be addressed and improved. We summarise recent primary studies in Table 9.

**Table 9.** Specific population health primary studies addressing PPI.

| Population Health Specific PPI Challenge Area | Study | Aspects of note |
| --- | --- | --- |
| Data-driven | Johnson et al (42) | • There is little guidance on how to meaningfully involve the public in big data research.<br>• Involvement in big data research is significantly limited in comparison with other study designs.<br>• May be because common approaches to public involvement adopted in primary data research are not appropriate within big data analysis studies.<br>• The highly data driven discussions that underline this type of research can present a barrier to public involvement.<br>• There is now growing recognition that public involvement in big data research requires special considerations. |
| Data-driven | Hobbs et al (43) | Enhance public forum members' personal development in data-intensive health research through a personal development portfolio:<br>• Personal Profile - Personal details including education, qualifications and employment<br>• Relevant Experience - Volunteering and personal experience<br>• Training Record - Training events attended and events where been trainer or facilitator<br>• Personal statement - Overall description of skills and |
|  |  | experience they may have gained from involvement activities<br>• Involvement activities - Summary of each activity, skills and experience gained, evidence such as certificates or feedback and personal reflections on their involvement in this activity<br>• References - Details of relevant individuals and how known to the public contributor. |
| Data-driven | 'Consensus Statement on Public Involvement and Engagement with Data Intensive Health Research'<sup>44</sup> | Key Principles for Public Involvement and Engagement in Data-Intensive Health Research –<br>1. Have institutional buy-in<br>2. Have clarity of purpose<br>3. Be transparent<br>4. Have two-way communication<br>5. Be inclusive and accessible to broad publics<br>6. Be ongoing<br>7. Be designed to produce impact<br>8. Be evaluated. |
| Complexity | Van Voorn et al (45) | • Involving patients in health economic research will require a serious investment of time and money for patients to get to a level at which they can contribute.<br>• Patients need to be able to 'rise above' their condition - to find an interest in the material itself and have an objective view.<br>• Proper selection procedures will have to be developed. |
| Representation & data-driven | Jewell et al (46) | Report on the setting up of a service user and carer advisory group supporting data linkage in mental health research.<br>• The general public feel that the complexities of data linkage research may be difficult to explain in lay terms and that patients and the public have limited knowledge about data, anonymisation, aggregation, and the regulations surrounding these.<br>• Training sessions were set up for all new group members. Training sought to provide members with information about data linkage, including the information governance procedures in place to protect the personal data of service users. |

The specific aspect of longer-term duration that is often typical of population health studies is best illustrated through the examination of existing longitudinal studies. Longitudinal studies involve repeated observations of the same subjects, allowing researchers to analyse change at the individual level. Such studies typically last decades, such as the 1970 British Cohort Study (47) or the Medical Research Council National Survey of Health and Development (48) which started in 1946.

Considering involvement in longitudinal studies, one approach is that used by the ALSPAC study (49). Based at the University of Bristol, the Avon Longitudinal Study of Parents and Children (ALSPAC), also known as Children of the 90s, is a world-leading birth cohort study. One of the governance aspects of the study is the original cohort advisory panel (OCAP) which is made up of more than 30 study participants who meet bi-monthly to provide insights and advice on study design, methodology and acceptability for participants. The group has been running since 2006.

The main aims of the OCAP group are:

- To represent the cohort of original study children
- To review study documentation and provide feedback to CO90s staff
- To represent and convey participants’ opinions about planned research exercises.

These additional sources suggest that certain solutions identified in the reviews, such as good communication and tailored training, are even more vital to PPI in population health research. One thing that emerges strongly from these studies is the idea that PPI selection and recruitment for population health research projects needs to be very carefully considered.

### UK Standards

The UK Standards proved to be a coherent framework for capturing solutions and no solution was offered that did not fit in to one of the six categories. It was, however, notable that two standards were less discussed than others: Governance and Impact. Capturing, measuring and illustrating the impact of PPI within the entire lifespan of a project is an issue that has not yet been resolved but is currently being addressed by various organisations. The absence of Governance may be a result of language use, as some attributes of Working Together were relevant in terms of this standard but were not couched in terms of Governance specifically. It was also interesting to see that Communications is a UK Standard separate from Working Together, as it was something that could be seen to be an integral part of Working Together.

### What is missing?

The solutions were matched against the problems identified as population health challenges. Not all challenges were matched by solutions as shown in the table below (Table 10). Challenges not matched to solutions are in bold and have been allocated to an appropriate UK Standard. By combining the additional challenges to those matched against solutions we can anticipate producing a more comprehensive list of things to address to help ensure that PPI within a project is both more effective in itself, and also helps to achieve better outcomes for the research.

**Table 10.**
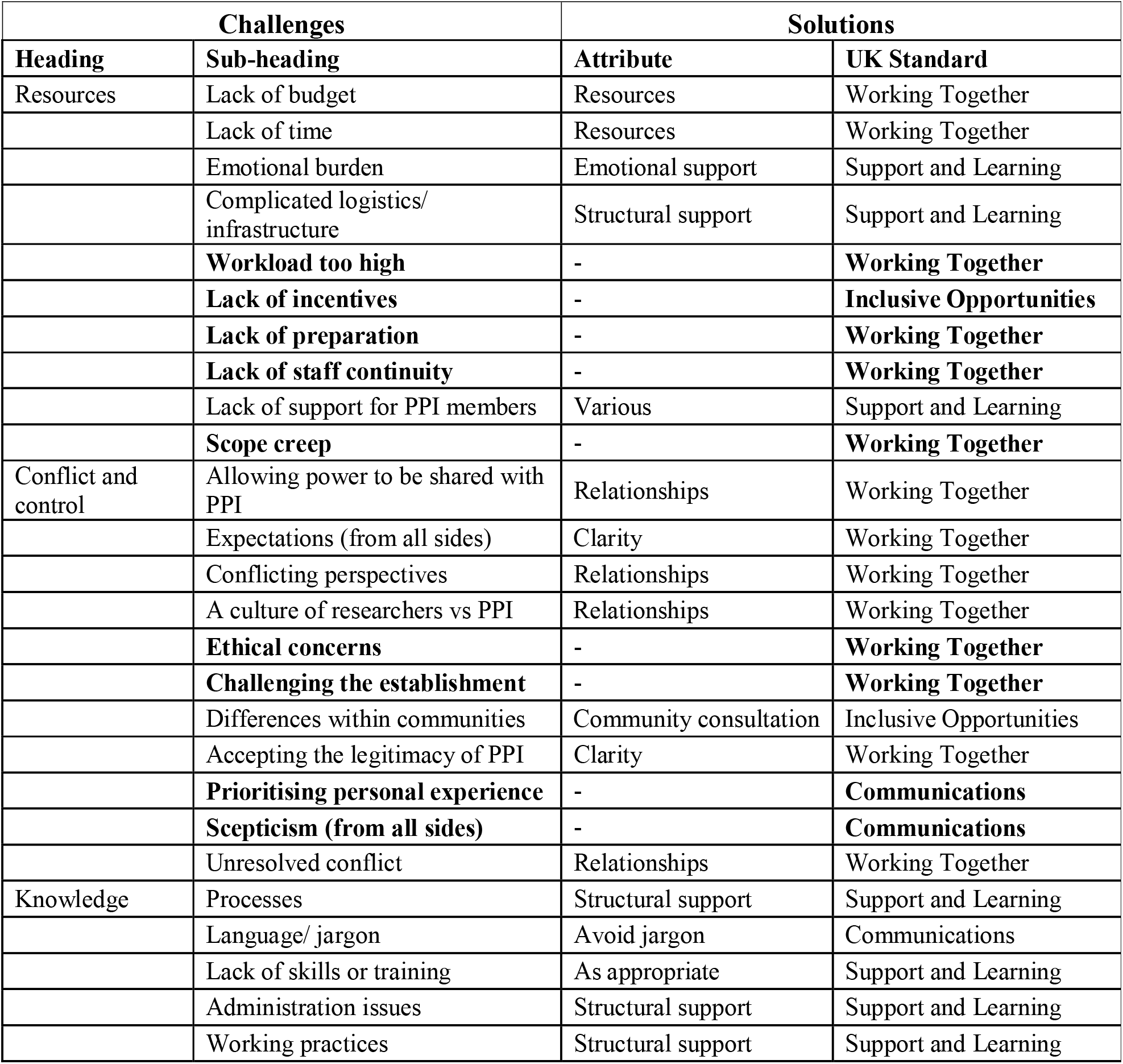
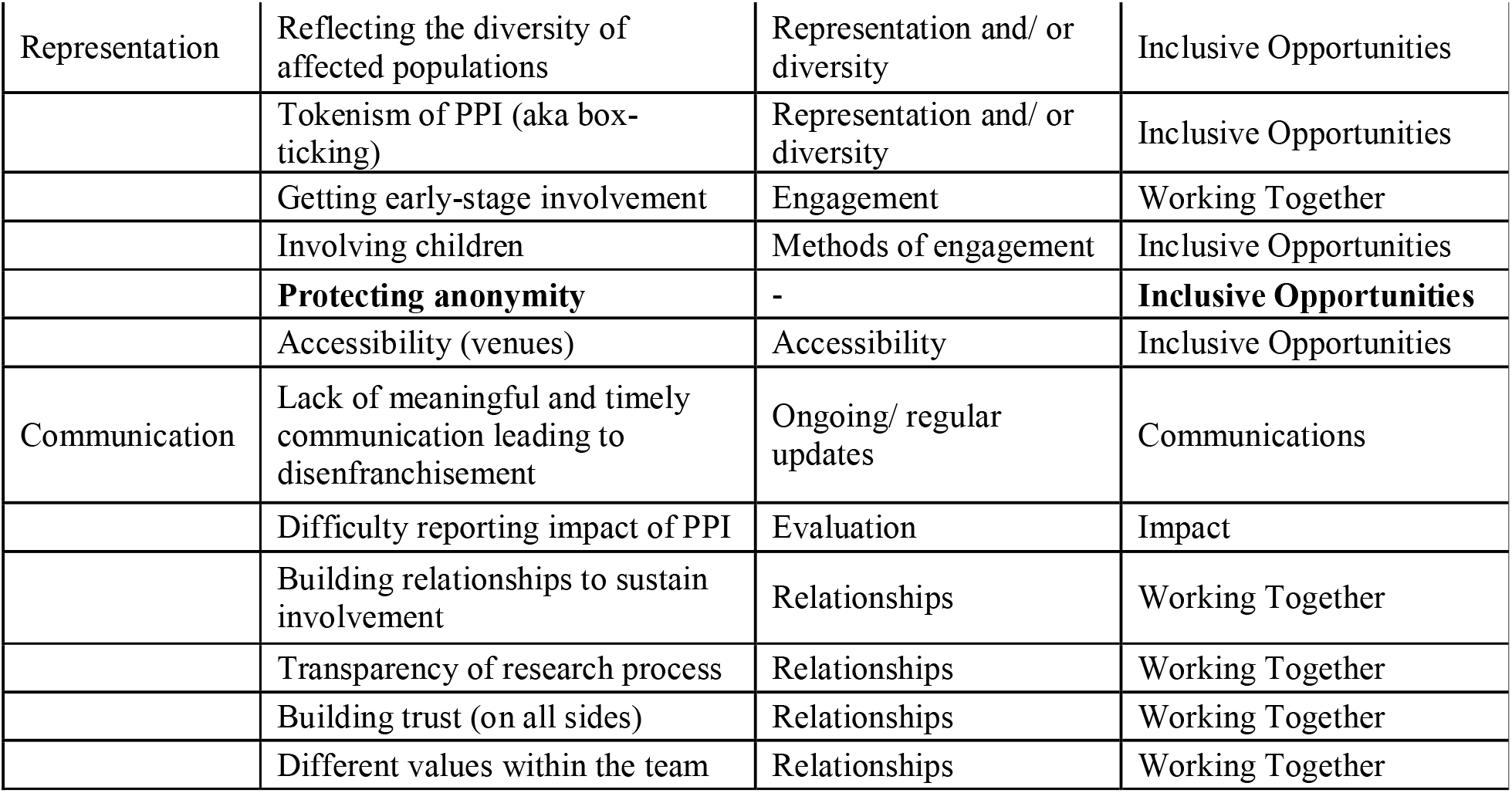
Challenges of facilitating good PPI in health research matched against solutions.

### Impact - Evaluation of PPI

Most studies focused on the impact of PPI activity on participants, researchers or the research itself – rather than setting out to formally assess what works to make PPI activity successful. For example, the Public and Patient Engagement Evaluation Tool (PPEET), developed by McMaster University in Canada, provides three questionnaires that examine involvement from the point of view of the participant, the project or the organisation (50). Although there are some tools to assess impact in PPI they tend to focus on certain aspects, for example the GRIPP2 tool assesses reporting of PPI (51).

This relative lack of scrutiny makes it difficult to assess actions that a project could or should undertake to help ensure best practice in PPI.

The evaluation guidance provided on the National Institute for Health Research (NIHR) website suggests the following categories of evaluation:

- Impact log – a simple method of recording outcomes
- ‘Cube’ framework – used to evaluate the process or quality of involvement
- Public Involvement Impact Assessment Framework (PiiAF) – more comprehensive method consisting of two parts 1) planning involvement in a research project, 2) designing a plan to evaluate the impact of involvement
- Realist evaluation – identifies what works for whom in what circumstances and what respects, and how (52).

Moreover, there is much still to be decided about what impact may be reasonably expected to be seen. Brett et al (53) notes particularly the lack of any evidence of any financial analysis and Jones et al (54) suggests that the use of contemporaneous real time data concerning PPI within surgical trials, currently lacking, could be made use of. Furthermore, ‘[T]he impact of involvement will always be somewhat unpredictable, because at the start of any project researchers ‘don’t know what they don’t know’—they do not know precisely what problems they might anticipate, until the patients/public tell them…. One of the most important contextual factors that influence the outcome of involvement is the researchers themselves, in particular the skills, knowledge, values and assumptions they start with. They are often the ‘subjects’ who experience the impact of involvement. For this reason, the answer to the question ‘Is involvement worth doing?’ will always be ‘It depends’ (55).

One further point of consideration is that it could be considered that the aspirational end point of PPI would be that any involvement would become so integral to the project that it would be difficult to unpick whose contribution had led to an impact or outcome not originally anticipated.

Developing new tools for use in population health research

Based the synthesis of reviews, a guidance framework for use in health research projects including PPI and based on the UK Standards was developed (Supporting Information 4). It combines the solutions and outstanding challenges taken from the review of reviews and aims to provide a useful tool for researchers to enhance and improve their PPI activity within population health and general health research.

From this framework, and with additional information taken from the studies detailed in Table 9, the following table (Table 11) has been developed which looks specifically at actions which could help mitigate the key challenges of PPI in population health research.

**Table 11.**
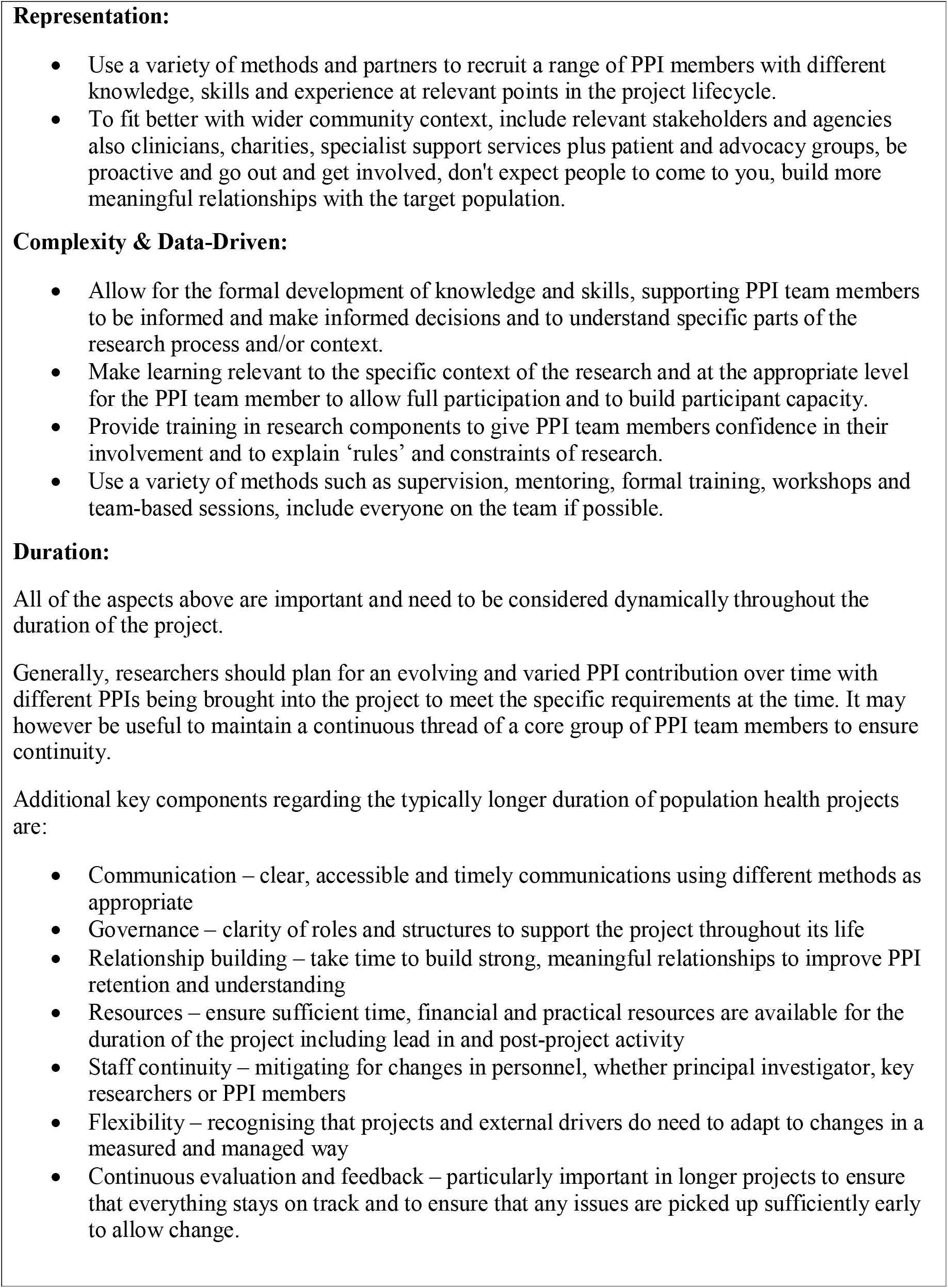
Recommended actions to address PPI in population health research.

What these tools (Table 11 and Supporting Information 4) do not provide is:

- A comprehensive tick list style approach
- Everything you could possibly consider – only what is in the reviews
- An indication of which actions are more important than others.

Research projects are different enough from each other that generic instructions will never suit every project. These tools therefore aim to provide pointers on important areas to consider without being directive about how that might look in a particular project.

### Strengths and limitations of the review of reviews

The review of reviews was carried out using systematic processes and following production of an a priori protocol. Not all data were however complete for all reviews and there was a wide variety within the reviews that did report data. For example,

- The number of studies reported in each review varied from 4 (37) to 251 (35)
- Years searched ranged from time periods defined by the previous decade (40) to those that searched back to the inception of the databases searched (26)
- Geography also varied but, of those reviews which gave details of geographical settings, the vast majority of the studies were from the UK (n = 292), followed by the USA (n = 95) and then other areas: Canada (n = 38), Europe (n = 29), Australia (n = 25), and other countries or multiple site studies (n = 17).

The reviews covered a range of diagnostic areas ranging from generic health and social care (14) or clinical trials (41) to condition specific areas such as diabetes (33) or palliative care (17).

Interestingly there were few reviews based on demographic groups who are generally acknowledged to be under-represented in healthcare decision making:

- There was one review for Black and Asian Minority Ethnic communities (15) and the geography of the studies included were mainly in the United States.
- There was one review for Older People (20) which covered nine qualitative articles. Arguably studies around dementia and palliative care may be relevant to this demographic but that cannot be assumed.
- There were three reviews for Children and Young People – all of which had a specific focus rather than looking at the involvement of Children and Young People in PPI more generally:
  ▪ Children and Families in Pediatric Health Research (19)
  ▪ Disabled children (21)
  ▪ Paediatric Intensive Care (37).

On the positive side, Malterud et al (56) however noted the usefulness of ‘two articles [which] describe in detail how individuals with limited literacy abilities can be supported to analyse and communicate such processes’.

## Conclusions

There are several important areas of PPI activity that require further research. With regards to Population Health research, there remain gaps in knowledge about how to ensure key specific attributes of this type of research are addressed for PPI team members – particularly around how to deal with complexity and the data-driven nature of the research. Looking at the UK Standards when mapped against the findings, it is clear that there is a lack of current research or clarity around Governance and Impact. There could also be more research done about PPI with under-represented groups. The new tools produced from the synthesis are designed to help population health researchers to facilitate better PPI and in turn to conduct better research.

## Data Availability

All relevant data are within the manuscript and its Supporting Information files.

## Acknowledgements

This review of reviews was discussed with the Centre for Population Health Patient and Public Involvement Advisory Group which meets quarterly to help set the strategic direction for PPI within the Centre. The draft review was read and commented on several times throughout its development by Dr. Helen Davies and Sarah Peddle – two of the PPI advisory group members. The authors would like to thank the group, and particularly Helen and Sarah, for their valuable input. The authors would also like to thank Kiara Jackson for providing input to the quality assessment section and Dr. Mayara Silveira Bianchim for input in the final version.

## Abbreviations

ALSPAC: Avon Longitudinal Study of Parents and Children
CO90s: Children of the 90s
CPH: National **Centre for Population Health** and Wellbeing Research
HCRW: Health and Care Research Wales
HE: Health Economics
HTA: Health Technology Assessment
IKT: Integrated knowledge translation
OCAP: Original Cohort Advisory Panel
PPEET: Public and Patient Engagement Evaluation Tool
PPE: Patient and Public Engagement/ Public and Patient Engagement
PPI: Patient and Public Involvement / Public and Patient Involvement

## Declarations

### Authors’ contributions

JN and SV designed the review of reviews. SV undertook data processing and JN provided advice, oversight and checked data processing and validity. Both authors developed the manuscript and approved the final version.

## Supporting Information

**S1 File. Search Strategy**. Full search strategy used.

**S2 Table. Quality Assessments**. Results of the quality Assessment.

**S3 Table. Full list of included studies**. A description of all included studies is provided as a table.

**S4 Table. PPI Guidance Framework**. Full PPI Guidance Framework.

## Footnotes

1 When a project outgrows its original remit without any additional resources being available.

## References

1. What is public involvement in research? Health Research Authority. 2020 Dec 16 [cited 2021 April 15]. Available from: https://www.hra.nhs.uk/planning-and-improving-research/best-practice/public-involvement/

2. Manafo E, Petermann L, Mason-Lai P, Vandall-Walker V. Patient engagement in Canada: a scoping review of the ‘how’ and ‘what’ of patient engagement in health research. Health Res Policy Syst. 2018 Mar 14;16(1):24. doi: 10.1186/s12961-018-0296-y.

3. Holmes J. What does improving population health really mean? The King’s Fund. 2022 July 21 [cited 2022 July 21]. Available from: https://www.kingsfund.org.uk/publications/what-does-improving-population-health-mean

4. WHO. Public Health Services [cited 2021 September 09]. Available from: https://www.euro.who.int/en/health-topics/Health-systems/public-health-services

5. Diez-Roux AV. On the Distinction—or Lack of Distinction—Between Population Health and Public Health. Am J Public Health. 2016 April; 106(4): 619–620. doi: 10.2105/AJPH.2016.303097

6. Ali H. I am not ‘hard to reach’. UpRising. 2020 April 9 [cited 2021 November 7]. Available from: https://www.uprising.org.uk/news/i-am-not-hard-reach

7. Improving inclusion of under-served groups in clinical research: Guidance from INCLUDE project. National Institute for Health and Care Research, 2020 August 7 [cited 2021 November 7]. Available from: https://www.nihr.ac.uk/documents/improving-inclusion-of-under-served-groups-in-clinical-research-guidance-from-include-project/25435

8. Wildridge, V & Bell, L 2002, ‘How clip became eclipse: A mnemonic to assist in searching for health policy/management information’, Health Information & Libraries Journal, vol. 19, no. 2, pp. 113–115.

9. AMSTAR 2 – The new and improved AMSTAR. AMSTAR. 2021 [cited 2021 April 15]. Available from: https://amstar.ca/Amstar-2.php

10. CASP Checklists. CASP [cited April 15]. Available from: https://casp-uk.net/casp-tools-checklists/

11. Iliffe S, Wilcock J, Drennan V, et al. Changing practice in dementia care in the community: developing and testing evidence-based interventions, from timely diagnosis to end of life (EVIDEM). Southampton (UK): NIHR Journals Library; 2015 Apr. (Programme Grants for Applied Research, No. 3.3.) Appendix 65, Chapter 5: Five main stages in framework analysis. [cited December 12]. Available from: https://www.ncbi.nlm.nih.gov/books/NBK286110/

12. The UK Standards: Setting the scene. UK Standards for Public Involvement. [cited 2021 April 09]. Available from: https://sites.google.com/nihr.ac.uk/pi-standards/standards/setting-the-scene

13. Boote J, Telford R, Cooper C. Consumer involvement in health research: a review and research agenda. Health Policy. 2002 Aug;61(2):213–36. doi: 10.1016/s0168-8510(01)00214-7. PMID: 12088893.

14. Brett J, Staniszewska S, Mockford C, Herron-Marx S, Hughes J, et al. A systematic review of the impact of patient and public involvement on service users, researchers and communities. Patient. 2014;7(4):387–95. doi: 10.1007/s40271-014-0065-0. PMID: 25034612.

15. Dawson S, Campbell SM, Giles SJ, Morris RL, Cheraghi-Sohi S. Black and minority ethnic group involvement in health and social care research: A systematic review. Health Expect. 2018 Feb;21(1):3–22. doi: 10.1111/hex.12597. Epub 2017 Aug 15. PMID: 28812330; PMCID: PMC5750731.

16. Zych MM, Berta WB, Gagliardi AR. Conceptualising the initiation of researcher and research user partnerships: a meta-narrative review. Health Res Policy Sys 18, 24 (2020). doi.org/10.1186/s12961-020-0536-9

17. Scholz B, Bevan A, Georgousopoulou E, Collier A, Mitchell I. Consumer and carer leadership in palliative care academia and practice: A systematic review with narrative synthesis. Palliat Med. 2019 Sep;33(8):959–968. doi: 10.1177/0269216319854012. Epub 2019 Jun 14. PMID: 31199194.

18. Pii KH, Schou LH, Piil K, Jarden M. Current trends in patient and public involvement in cancer research: A systematic review. Health Expect. 2019 Feb;22(1):3–20. doi: 10.1111/hex.12841. Epub 2018 Oct 30. PMID: 30378234; PMCID: PMC6351419.

19. Flynn R, Walton S, Scott SD. Engaging children and families in pediatric Health Research: a scoping review. Res Involv Engagem 5, 32 (2019). doi.org/10.1186/s40900-019-0168-9

20. Baldwin JN, Napier S, Neville S, Wright-St Clair VA. Impacts of older people’s patient and public involvement in health and social care research: a systematic review. Age Ageing. 2018 Nov 1;47(6):801–809. doi: 10.1093/ageing/afy092. PMID: 29939208.

21. Bailey S, Boddy K, Briscoe S, Morris C. Involving disabled children and young people as partners in research: a systematic review. Child Care Health Dev. 2015 Jul;41(4):505–14. doi: 10.1111/cch.12197. Epub 2014 Oct 16. PMID: 25323964.

22. Brett J, Staniszewska S, Mockford C, Herron-Marx S, Hughes J, et al. Mapping the impact of patient and public involvement on health and social care research: a systematic review. Health Expect. 2014 Oct;17(5):637–50. doi: 10.1111/j.1369-7625.2012.00795.x. Epub 2012 Jul 19. PMID: 22809132; PMCID: PMC5060910.

23. Howard WS, Brainard J, Loke Y. et al. Patient and public involvement in health literacy interventions: a mapping review. Res Involv Engagem 3, 31 (2017). doi.org/10.1186/s40900-017-0081-z

24. Price A, Albarqouni L, Kirkpatrick J, Clarke M, Liew SM, Roberts N, et al. Patient and public involvement in the design of clinical trials: An overview of systematic reviews. J Eval Clin Pract. 2018 Feb;24(1):240–253. doi: 10.1111/jep.12805. Epub 2017 Oct 27. PMID: 29076631.

25. Bethell J, Commisso E, Rostad HM, Puts M, Babineau J, Grinbergs-Saull A, et al. Patient engagement in research related to dementia: A scoping review. Dementia (London). 2018 Nov;17(8):944–975. doi: 10.1177/1471301218789292. PMID: 30373460.

26. Domecq JP, Prutsky G, Elraiyah T, et al. Patient engagement in research: a systematic review. BMC Health Serv Res 14, 89 (2014). doi.org/10.1186/1472-6963-14-89

27. Boote J, Baird W, Beecroft C. Public involvement at the design stage of primary health research: a narrative review of case examples. Health Policy. 2010 Apr;95(1):10–23. doi: 10.1016/j.healthpol.2009.11.007. Epub 2009 Dec 5. PMID: 19963299.

28. Nunn JS, Tiller J, Fransquet P, Lacaze P. (2019). Public Involvement in Global Genomics Research: A Scoping Review. Frontiers in public health, 7, 79. doi.org/10.3389/fpubh.2019.00079

29. Sangill C, Buus N, Hybholt L, Berring LL. (2019). Service user’s actual involvement in mental health research practices: A scoping review. International Journal of Mental Health Nursing, 28(4), 798–815. doi.org/10.1111/inm.12594

30. Fergusson D, Monfaredi Z, Pussegoda K, et al. The prevalence of patient engagement in published trials: a systematic review. Res Involv Engagem 4, 17 (2018). doi.org/10.1186/s40900-018-0099-x

31. Jagosh J, Macaulay AC, Pluye P, Salsberg J, Bush PL, Henderson J, et al. Uncovering the benefits of participatory research: implications of a realist review for health research and practice. Milbank Q. 2012 Jun;90(2):311–46. doi: 10.1111/j.1468-0009.2012.00665.x. PMID: 22709390; PMCID: PMC3460206.

32. Camden C, Shikako-Thomas K, Nguyen T, Graham E, Thomas A, Sprung J, et al. Engaging stakeholders in rehabilitation research: a scoping review of strategies used in partnerships and evaluation of impacts. Disabil Rehabil. 2015;37(15):1390–400. doi: 10.3109/09638288.2014.963705. Epub 2014 Sep 22. PMID: 25243763.

33. Harris J, Haltbakk J, Dunning T, et al. How patient and community involvement in diabetes research influences health outcomes: A realist review. Health Expect. 2019;22(5):907–920. doi:10.1111/hex.12935

34. Baines RL, Regan de Bere S. Optimizing patient and public involvement (PPI): Identifying its “essential” and “desirable” principles using a systematic review and modified Delphi methodology. Health Expect. 2018 Feb;21(1):327–335. doi: 10.1111/hex.12618. Epub 2017 Sep 19. PMID: 28929554; PMCID: PMC5750770.

35. Vaughn LM, Whetstone C, Boards A, Busch MD, Magnusson M, Määttä S. Partnering with insiders: A review of peer models across community-engaged research, education and social care. Health Soc Care Community. 2018 Nov;26(6):769–786. doi: 10.1111/hsc.12562. Epub 2018 Mar 7. PMID: 29512217.

36. Chambers E, Gardiner C, Thompson J, Seymour J. Patient and carer involvement in palliative care research: An integrative qualitative evidence synthesis review. Palliat Med. 2019 Sep;33(8):969–984. doi: 10.1177/0269216319858247. Epub 2019 Jun 28. PMID: 31250702; PMCID: PMC6691598.

37. Menzies JC, Morris KP, Duncan HP. et al. Patient and public involvement in Paediatric Intensive Care research: considerations, challenges and facilitating factors. Res Involv Engagem 2, 32 (2016). doi.org/10.1186/s40900-016-0046-7

38. Shippee ND, Domecq GJP, Prutsky LGJ, Wang Z, Elraiyah TA, Nabhan M, et al. Patient and service user engagement in research: a systematic review and synthesized framework. Health Expect. 2015 Oct;18(5):1151–66. doi: 10.1111/hex.12090. Epub 2013 Jun 3. PMID: 23731468; PMCID: PMC5060820.

39. Dawson S, Campbell SM, Giles SJ, Morris RL, Cheraghi-Sohi S. Black and minority ethnic group involvement in health and social care research: A systematic review. Health Expect. 2018 Feb;21(1):3–22. doi: 10.1111/hex.12597. Epub 2017 Aug 15. PMID: 28812330; PMCID: PMC5750731.

40. Pii KH, Schou LH, Piil K, Jarden M. Current trends in patient and public involvement in cancer research: A systematic review. Health Expect. 2019 Feb;22(1):3–20. doi: 10.1111/hex.12841. Epub 2018 Oct 30. PMID: 30378234; PMCID: PMC6351419.

41. Crocker JC, Ricci-Cabello I, Parker A, Hirst JA, Chant A, Petit-Zeman S, et al. Impact of patient and public involvement on enrolment and retention in clinical trials: systematic review and meta-analysis BMJ 2018; 363 :k4738 doi:10.1136/bmj.k4738

42. Johnson H, Davies JM, Leniz J, Chukwusa E, Markham S, Sleeman KE. Opportunities for public involvement in big data research in palliative and end-of-life care. Palliative Medicine. 2021;35(9):1724–1726. doi:10.1177/02692163211002101

43. Hobbs G, Tully MP. Realist evaluation of public engagement and involvement in data-intensive health research. Res Involv Engagem 6, 37 (2020). doi.org/10.1186/s40900-020-00215-4

44. Aitken M, Tully MP, Porteous C, Denegri S, Cunningham-Burley S, Banner N, et al. (2020) “Consensus Statement on Public Involvement and Engagement with Data-Intensive Health Research”, International Journal of Population Data Science, 4(1). doi: 10.23889/ijpds.v4i1.586.

45. van Voorn GA, Vemer P, Hamerlijnck D, et al. The Missing Stakeholder Group: Why Patients Should be Involved in Health Economic Modelling. Appl Health Econ Health Policy. 2016;14(2):129–133. doi:10.1007/s40258-015-0200-7

46. Jewell A, Pritchard M, Barret, K. et al. The Maudsley Biomedical Research Centre (BRC) data linkage service user and carer advisory group: creating and sustaining a successful patient and public involvement group to guide research in a complex area. Res Involv Engagem 5, 20 (2019). doi.org/10.1186/s40900-019-0152-4

47. 1970 British Cohort Study. Centre for Longitudinal Studies. [cited 2021 November 7]. Available from: https://cls.ucl.ac.uk/cls-studies/1970-british-cohort-study/

48. National Survey of Health and Development. Medical Research Council. [cited 2021 November 7]. Available from: https://www.nshd.mrc.ac.uk/

49. Avon Longitudinal Study of Parents and Children. University of Bristol. [cited 2021 November 7]. Available from: http://www.bristol.ac.uk/alspac/about/

50. Public and Patient Engagement Evaluation Tool. Faculty of Health Sciences. [cited 2021 November 25]. Available from: https://ppe.mcmaster.ca/our-products/public-patient-engagement-evaluation-tool

51. Staniszewska S, Brett J, Simera I, Seers K, Mockford C, Goodlad S et al. GRIPP2 reporting checklists: tools to improve reporting of patient and public involvement in research BMJ 2017; 358 :j3453 doi:10.1136/bmj.j3453

52. Kok M (2018) Guidance Document: Evaluating public involvement in research. UWE Bristol. [UWE Bristol e-prints repository]

53. Brett J, Staniszewska S, Mockford C, Seers K, Herron-Marx S, Bayliss H. The PIRICOM study: a systematic review of the conceptualisation, measurement, impact and outcomes of patients and public involvement in health and social care research: University of Warwick; 2010.

54. Jones EL, Williams-Yesson BA, Hackett RC, Staniszewska SH, Evans D, Francis NK. Quality of reporting on patient and public involvement within surgical research: a systematic review. Ann Surg. 2015 Feb;261(2):243–50. doi: 10.1097/SLA.0000000000000768. PMID: 24950279.

55. Staley, K. ‘Is it worth doing?’ Measuring the impact of patient and public involvement in research. Res Involv Engagem 1, 6 (2015). doi.org/10.1186/s40900-015-0008-5

56. Malterud K, Elvbakken KT. Patients participating as co-researchers in health research: A systematic review of outcomes and experiences. Scand J Public Health. 2020 Aug;48(6):617–628. doi: 10.1177/1403494819863514. Epub 2019 Jul 18. PMID: 31319762.

57. Jenkins M. Evaluation of methodological search filters--a review. Health Info Libr J. 2004 Sep;21(3):148–63. doi: 10.1111/j.1471-1842.2004.00511.x.

58. Manafò E, Petermann L, Vandall-Walker V, Mason-Lai P (2018) Patient and public engagement in priority setting: A systematic rapid review of the literature. PLoS ONE 13(3):e0193579. doi.org/10.1371/journal.pone.0193579.

59. Ocloo J, Garfield S, Dawson S, et al. Exploring the theory, barriers and enablers for patient and public involvement across health, social care and patient safety: a protocol for a systematic review of reviews. BMJ Open 2017;7:e018426. doi:10.1136/bmjopen-2017-018426

60. Rogers M, Bethel A, Boddy K. Development and testing of a MEDLINE search filter for identifying patient and public involvement in health research. Health Information & Libraries Journal, 34, pp. 125–133

