## Supplemental file 1. for "A review of reviews exploring patient and public involvement in population health research"

### Search strategy

One issue with research on patient and public involvement in population health research is the profusion of terms that may or may not mean a si

milar thing. After examining a sample of previous studies (Jenkins^57^, Manafo^58^, Ocloo^59^, Rogers^60^) the following overall list of potential terms was established.

Regarding involvement:

- PPI, Public and patient involvement, PPE, Public and patient engagement, community engagement, community involvement, active participation, involvement, collaboration, engagement, partnership, consultation, participation, user-led, consumer or patient panel, advisory board/ group/panel, building relationships, participatory research.

Regarding public:

- Community groups, community representatives, public, patient, carer, consumer, citizen, lay, layperson, service user, stakeholder, family, relative, survivor.

Regarding population health research:

- Population Health research, health services research, health care research, social care research, public health research, mental health research.

Regarding type of study:

- Systematic review, narrative review, bibliometric review, meta-analysis.

**PubMed Search**

(((PPI OR Public and patient involvement OR PPE OR Public and patient engagement OR Community engagement OR Community involvement OR active participation OR involvement OR collaboration OR engagement OR partnership OR consultation OR participation OR user-led OR consumer OR patient panel OR advisory board OR advisory group OR advisory panel OR Building relationships OR participatory research AND ((journalarticle[Filter] OR meta-analysis[Filter] OR systematicreviews[Filter]) AND (y_10[Filter]) AND (english[Filter]))) AND (community groups OR community representatives OR public OR patient OR carer OR consumer OR citizen OR lay OR layperson OR service user OR stakeholder OR family OR relative OR survivor AND ((journalarticle[Filter] OR meta-analysis[Filter] OR systematicreviews[Filter]) AND (y_10[Filter]) AND (English[Filter])))) AND (Population Health Research[Title/Abstract] OR health services research[Title/Abstract] OR health care research[Title/Abstract] OR social care research[Title/Abstract] OR public health research[Title/Abstract] OR mental health research[Title/Abstract] AND ((journalarticle[Filter] OR meta-analysis[Filter] OR systematicreviews[Filter]) AND (y_10[Filter]) AND (English[Filter]))) AND ((journalarticle[Filter] OR meta-analysis[Filter] OR systematicreviews[Filter]) AND (y_10[Filter]) AND (English[Filter]))) AND (Systematic review OR narrative review OR bibliometric review OR meta-analysis AND ((journalarticle[Filter] OR meta-analysis[Filter] OR systematicreviews[Filter]) AND (y_10[Filter]) AND (English[Filter])))

united kingdom or uk or britain or scotland or england or wales or northern ireland OR Europe* OR "New Zealand" OR Canada

Limiters - Scholarly (Peer Reviewed) Journals; Date of Publication: 20100101-20201231; English Language; Human
