## Supplemental Table 1 for "A review of reviews exploring patient and public involvement in population health research"

### **Quality Assessment**

| **Question ->** | **Did the review address a clearly focused question?** | **Did the authors look for the right type of papers?** | **Do you think all the important, relevant studies were included?** | **Did the review’s authors do enough to assess quality of the included studies?** | **If the results of the review have been combined, was it reasonable to do so?** | **How precise are the results?** | **Can the results be applied to the local population?** | **Were all important outcomes considered?** | **Are the benefits worth the harms and costs?** |
| --- | --- | --- | --- | --- | --- | --- | --- | --- | --- |
| **Paper** | **Agreed** | **Agreed** | **Agreed** | **Agreed** | **Agreed** | **Agreed** | **Agreed** | **Agreed** | **Agreed** |
| **Bailey** | Y | Y | Y | Y | Y | CT | Y | Y | Y |
| **Baines** | Y | Y | Y | Y | Y | % | Y | Y | Y |
| **Baldwin** | Y | Y | Y | Y | Y | CT | Y | Y | Y |
| **Bethell** | Y | Y | N | N | Y | CT | Y | Y | Y |
| **Boote** | Y | Y | N | N | Y | CT | Y | Y | Y |
| **Brett^47^** | Y | Y | Y | Y | Y | CT | Y | Y | Y |
| **Brett^9^** | Y | Y | Y | Y | Y | CT | Y | Y | Y |
| **Brett^17^** | Y | Y | Y | Y | Y | CT | Y | Y | Y |
| **Camden** | Y | Y | Y | N | Y | CT | Y | Y | Y |
| **Chambers** | Y | Y | Y | Y | Y | CT | Y | Y | Y |
| **Crocker** | Y | Y | N | N | Y | % | Y | Y | Y |
| **Dawson** | Y | Y | N | Y | Y | CT | Y | Y | Y |
| **Domecq** | Y | Y | Y | Y | Y | CT | Y | Y | Y |
| **Fergusson** | Y | Y | N | N | Y | % | Y | Y | Y |
| **Flynn** | Y | Y | N | N | Y | CT | Y | Y | Y |
| **Harris** | Y | Y | Y | N | Y | CT | Y | Y | Y |
| **Jagosh** | Y | Y | N | N | Y | CT | Y | y | Y |
| **Jones** | Y | Y | Y | Y | Y | CT | Y | Y | Y |
| **Malterud** | Y | Y | N | Y | Y | CT | Y | Y | Y |
| **Manafo** | Y | Y | N | Y | Y | CT | Y | Y | Y |
| **Menzies** | Y | Y | N | Y | Y | CT | Y | Y | Y |
| **Miah** | Y | Y | Y | N | Y | CT | Y | Y | Y |
| **Nunn** | Y | Y | N | N | Y | % | Y | Y | Y |
| **Pii** | Y | Y | CT | Y | Y | CT | Y | Y | Y |
| **Price** | Y | Y | Y | Y | Y | CT | Y | Y | Y |
| **Sangill** | Y | Y | Y | Y | Y | CT | Y | Y | Y |
| **Scholz** | Y | Y | Y | Y | Y | CT | Y | Y | Y |
| **Shippee** | Y | Y | Y | N | Y | CT | Y | Y | Y |
| **Vaughn** | Y | Y | N | N | Y | CT | Y | Y | Y |
| **Wilsher** | Y | Y | CT | N | Y | CT | Y | Y | Y |
| **Zych** | Y | Y | N | Y | Y | CT | Y | Y | Y |

**Key:**

Y = Yes

N = No

CT = Can’t tell

% = precision given a percentage score
