## Supplemental Table 2 for "A review of reviews exploring patient and public involvement in population health research"

### **Full list of included studies**

| **Lead author** | **Year published** | **Type of review** | **Area of focus** | **Types of included studies** |
| --- | --- | --- | --- | --- |
| Bailey, S | 2014 | Systematic | Disabled children and young people | Twenty-two studies were included: seven reviews, eight original research studies, three reports, three guidelines and one webpage. Nine examples of involvement were identified. Four documents had been produced in partnership with disabled children and young people. Topics included: subjective wellbeing, healthcare technologies, health service development, asthma and diabetes. |
| Baines, R. L. | 2017 | Systematic | Health and social care research | Twelve systematic reviews and 88 grey literature publications were reviewed exploring PPI in medicine, dentistry or nursing across one or more of research, regulation, health-care services and educational settings. |
| Baldwin, J | 2018 | Systematic | Older people in health and social care research | Nine qualitative articles included that formally evaluated inclusion of older people in PPI activities. Themes included: stroke research, care transitions, dementia, frailty, use of community services. Members were included in various stages of research. |
| Bethell, J | 2018 | Scoping | Dementia research | Fifty-four studies mainly qualitative but also including mixed methods, cluster-randomised and systematic reviews. Topic areas (within dementia) included: health literacy, barriers to participation, informed consent and priority setting. |
| Boote, J | 2010 | Narrative | Research design - primary health research | Seven studies were identified covering the following topics: breast-feeding, antiretroviral and nutrition interventions; paediatric resuscitation; exercise and cognitive behavioural therapy; hormone replacement therapy and breast cancer; stroke; and parents’ experiences of having a pre-term baby. Six studies reported public involvement in the development of a clinical trial, while one reported public involvement in the development of a mixed methods study. |
| Brett, J | 2012 | Systematic | Health and social care research | 66 studies, two were randomized controlled trials (RCTs), one was a pre-test/post-test study, one was a cohort, 46 were qualitative studies, nine were cross-sectional, five were case studies and two were case series. Topics included: diabetes, medical technology, healthcare policy and priority setting, mental health and asthma along with general health care questions. |
| Brett, J | 2014 | Systematic | Health and social care research | Sixty-five studies reported impacts of PPI on health and social care service users. Of these studies, 42 were qualitative studies, 12 were case studies, three were a case series, three were cross-sectional studies, and five were reviews of the evidence.  Thirty-five studies reported impacts of PPI on health and social care researchers. Twenty-six studies were qualitative, four were case studies, one was a case series, one was cross-sectional, and three reviewed the evidence.  Twenty-three studies reported impacts of PPI on the community involved in research (i.e. the wider patient group under research). Seventeen studies were qualitative studies, one was a cross-sectional study, four were case studies, and one was a review of the evidence. Topics included: general health care questions such as around partnerships and participation and specific topic areas such as: young drug users, breast cancer, primary care and schizophrenia. Also included studies on specific populations including: Aboriginal grandmothers and black Americans. |
| Brett, J | 2010 | Systematic | Health and social care research | Qualitative and quantitative studies involving small groups, communities, disease specific cohorts, population-based interventions and services, primary care, mental health, and population/societal level studies, health technology assessment, trials. PPI involvement in specific stages such as research prioritisation, ethics preparation, scientific review and data analysis. Specific involvement, e.g. older people and those with disabilities/ learning disabilities, immigrants, hard to reach populations. |
| Camden, C | 2014 | Scoping | Rehabilitation research (stakeholders) | Nineteen studies - qualitative and quantitative research studies, and opinion/reflection studies as long as they were describing strategies used in a specific study - and on topics (within rehabilitation) including: poverty and disability, mental health, knowledge translation, participation and occupational therapy. |
| Chambers, E | 2019 | Systematic | Palliative care | Ninety-three records covering 60 studies, all study designs included including grey literature. Topics (within palliative care) included: end of life care, chronic kidney disease, service improvement, carers and cancer research. |
| Crocker, J | 2018 | Systematic & meta-analysis | Clinical trials | Twenty-six studies included of non-randomised and randomised trials. Topics included: depression, podiatry related, specific cancer areas, pre-diabetes. Specific population groups included: African-American men, Gulf War veterans, new mothers, and specific age groups for men or women with or without specific diseases. |
| Dawson, S | 2017 | Systematic | Black and Minority Ethnic groups in health and social care research | Forty-five studies including qualitative, quantitative, mixed methods, case study and evaluations. Health topics included: cancer, hypertension, diabetes, brain injury, HIV and health promotion. Specific population groups included: African Americans, Chinese, Aboriginal, Latino and Afghan. |
| Domecq, J | 2014 | Systematic | Healthcare research | 142 studies - 8 systematic reviews, 7 randomized trials and 24 observational studies; the remaining majority (103) consisted of qualitative studies. Topics included: clinical trials, primary research design, cancer, children with cerebral palsy, breast cancer and palliative care. |
| Fergusson, D | 2018 | Systematic | Clinical trials – randomised and non-randomised | Twenty-three trials, of which 17 were randomized control trials, and six were non-randomized comparative trials. The trials engaged a range of 2-24 patients/ community representatives per study. Topics included: ethics training for co-researchers, chronic obstructive pulmonary disease, diabetes, nutrition and exercise, weight and healthy aging. Population groups involved included engagement of children and minorities. |
| Flynn, R | 2019 | Scoping | Children and families in pediatric health research | Seventeen articles including case studies, randomized control trials, qualitative design, mixed-methods, multi-methods, comparative effectiveness research and psychometric research. Topics (within pediatric health research) included: mental health, asthma, participation, cerebral palsy, research training and hospice care. |
| Harris, J | 2019 | Realist | Diabetes | Data extracted from 29 projects (total of 92 articles) with a focus on adults, families or communities including people at risk of diabetes as well as those already diagnosed based in clinical or community settings. |
| Jagosh, J | 2012 | Realist Lit Review | Health research and practice | Twenty-three participatory research partnerships described in 276 publications including peer reviewed and non-peer-reviewed publications and websites that reported on empirical research, activity descriptions, and authors’ reflections. Topics included: cardiovascular disease and diabetes prevention, weight loss, breast cancer prevention, cancer prevention, depression awareness and prevention, substance abuse and health promotion. |
| Jones, E | 2015 | Systematic | Surgical research | Eight full text articles were included describing a total of 489 patients. Surgical specialisms included: urology, musculo-skeletal, colorectal, ear nose and throat, and gynaecology. One additional study involving cancer patients from multiple surgical specialities and one systematic review study were also included. |
| Malterud, K | 2019 | Systematic | Co-researchers in health research | Seventeen primary studies. The studies represented patient groups with different health issues (mental health problems, learning disabilities, occupational injuries, cancer, osteoarthritis). The sample consisted of only qualitative studies (individual interviews, focus group studies, participatory action research, single case study, nominal study). |
| Manafo, E | 2018 | Scoping | Health research | Fifty-five records of which 44 =formal review and 11 = informal review/grey literature. Methodologies included: case study, experimental, quasi-experimental, non-experimental, qualitative, literature reviews and commentary. Topics included: participation and involvement in general health care, knowledge translation, clinical research, burn survivors and cardiovascular disease. |
| Menzies, JC | 2016 | Lit Rev | Paediatric intensive care | Four relevant studies. Three of the studies had consulted with parents of children who had been on paediatric intensive care but only one study had spoken directly to a child themselves. |
| Miah, J | 2019 | Scoping | Dementia research | Twenty studies on topics (within dementia research) including: screening, diagnosis and treatment, home care support, carer support, and individual cognitive stimulation therapy. Mainly qualitative. |
| Nunn, J. S. | 2019 | Scoping | Genomics | Thirty-two projects (or initiatives) on genomics included. Topics (within genomics) included: wellness, biobanking, clinical sequencing, rare diseases and undiagnosed diseases. |
| Pii, K | 2018 | Systematic | Cancer | Twenty-seven studies were included. The populations in the studies were defined in various ways, though most were disease-specific, while other populations were defined according to age or ethnicity. The majority of the studies focused on specific cancer diseases: breast cancer, lung cancer (n = 4), blood cancer, colorectal cancer, gynaecological cancer and bowel cancer. |
| Price, A | 2017 | Systematic | Clinical trials | Twenty-seven reviews included. Methodologies included: mixed methods, narrative and case examples, scoping, qualitative, and bibliometric. Topics (within clinical trials) included: disabled children, primary health research design, social care, engagement, rare diseases, older people, end of life, and cancer. |
| Sangill, C | 2019 | Scoping | Mental health | Thirty-two studies included. The methodologies of the studies ranged from five quantitative studies, out of which one had a randomized design, 24 qualitative studies, for example analyses of interviews and observations as well as case studies and pilot studies, and three mixed methods studies. There were 17 studies with a clear participatory/collaborative design. Topics (within mental health) included: peer research, collaboration, learning disability, detained psychiatric patients, and training. |
| Scholz, B | 2019 | Systematic | Palliative care | Eleven studies were included. Topics (within palliative care) included: student teaching in hospice, palliative and supportive care, mesothelioma, communication skills, and cancer care. |
| Shippee, N | 2013 | Systematic | Biomedical and health services research | 202 studies reported on. Topics included: cancer, mental health, participatory research, decision making, diabetes, community health interventions, chronic diseases, older people, and HIV prevention. |
| Vaughn, L. M. | 2018 | Review | Peer models, research, education and social care | 251 articles included. Articles were categorised as empirical, process/descriptive, and “about” peers. Topics included: cancer, smoking, mental health, environmental/occupational health, health promotion, and sexual health. |
| Wilsher, S. H. | 2017 | Mapping | Health literacy interventions | Twenty studies included. Most studies focused on patients with a range of chronic conditions and four on ethnic groups who had immigrated to America. Topics (within health literacy) included: self-care, chronic diseases, colorectal cancer screening among veterans, diabetes and asthma. Study formats included: randomised clinical trial, non-randomised clinical trial, and single arm. |
| Zych, M. M. | 2020 | Narrative | Partnership initiation - Healthcare and social sciences research | Seventeen reviews included. Methodologies included: theoretical review, scoping, systematic, realist, descriptive and narrative. Topics included: public health, health services, medicine, nursing, knowledge management, education and organisational management. |
