## Supplemental Table 3 for "A review of reviews exploring patient and public involvement in population health research"

### **PPI Guidance Framework**

| **Inclusive opportunities** | |
| --- | --- |
| **Heading** | **Aspects to consider** |
| Accessibility | Venues should be located for the ease of the participants, accessible and meetings should be timed appropriately and include communication aids, breaks and refreshments as appropriate for individual and collective needs. |
| Methods of engagement | Online technology could assist people to be included who are often excluded from traditional engagement e.g. those with illness, time poor, caring responsibilities. Especially when working with disabled children and young people, be flexible for different abilities and ages and offer choice, use variety of methods. |
| Incentives | Provide incentives for participants. |
| Anonymity | Ensure anonymity for participants if required or requested. |
| Representation and/or diversity | Use variety of methods and partners to recruit a range of participants, understand different motivations and gain insight into the community, view differing perspectives as valuable, recognise and address issues concerning diversity, avoid tokenism. |
| Community consultation | To fit better with wider community context, include relevant stakeholders and agencies also clinicians, charities, specialist support services plus patient and advocacy groups. Be proactive and go out and get involved, don't expect people to come to you, build more meaningful relationships with target population. |
| Safe environment | Consider whether a trusted adult or facilitator is useful. |
| Recruit well | Fit skills and experiences to the project, recruit through a variety of ways, need to be not just representative but also be able to be collaborative. |
| **Working together** | |
| **Heading** | **Aspects to consider** |
| Resources | Consider different aspects of resourcing such as budget/ funding, building in sufficient time to build relationships, communicate etc., consider using existing PPI resources or groups where available, plan into proposals, tailor to project. |
| Clarity | Ensure clarity of various aspects including: roles, expectations, structures. |
| Workload | Prevent workload getting too much. |
| Preparation | Prepare in advance as much as possible. |
| Staff continuity | Mitigate and prepare for staff turnover but avoid if possible. |
| Scope creep | Prevent or manage scope creep. |
| Relationships | Manage conflict, take time to build partnerships built on joint ownership, trust, respect and transparency, empower PPI members by sharing power and knowledge, explore risks together, consider capacity of PPI members. |
| Engagement | Conduct engagement early on, provide multiple and varied opportunities as appropriate. Always acknowledge contributions. |
| Flexibility | Acknowledge that confidence, personal circumstances and capacity may change over time, keep tasks flexible and include time for training and questions. Be flexible generally in attitude and approaches to the project. |
| Ethical concerns | Consider and address possible ethical concerns throughout and understand that they may not always be obvious. |
| Challenging the establishment | Have honest conversations about motivations of team members, including PPI and other stakeholders, on all sides. |
| **Support and learning** | |
| **Heading** | **Aspects to consider** |
| Practical support | Think about details e.g. childcare, food, location, transport, compensation, timings, and have strategies for when people are ill/ can’t take part. |
| Structural support | Make sure key project individuals support PPI, provide structures that support PPI, include relevant institutions such as charities, volunteer groups etc. |
| Formal knowledge | Encourage formal development of knowledge and skills, supporting participants to be informed and make informed decisions and to understand specific parts of the research process and/or context. |
| Learning as appropriate | Make learning relevant to the specific context of the research and at the appropriate level for the PPI member to allow full participation and to build participant capacity. |
| Research methods | Ensure access to training in research components to give confidence in their involvement and to explain ‘rules’ and constraints of research. |
| Emotional support | Recognise that experiences may be upsetting, provide safe spaces, provide consistent feedback and support, consider how to deal with anxiety. |
| Share knowledge | Acknowledge that knowledge and experience flow both ways and make ways to facilitate that flow. |
| Specific support | Ensure support specific to topic area and to their individual involvement. |
| Variety | Use a variety of methods such as supervision, mentoring, formal, workshops and team based, include everyone on the team if possible. |
| **Governance** | |
| **Heading** | **Aspects to consider** |
| Shared decision making | Create shared decision making (at every level), power and leadership. |
| **Communications** | |
| **Heading** | **Aspects to consider** |
| Have stakeholders lead groups | But be careful they include all groups in the discussion. |
| Ongoing/ regular updates | Provide regular update to contribute to motivation and engagement, and to foster satisfying partnerships. |
| Be open | Create space to voice concern/ open communication climate. |
| Avoid/ translate jargon | Ensure everyone understands and feels comfortable and confident to engage in meaningful dialogue. |
| Use different materials (not just written reports etc) | Ensure people with different levels of literacy can participate. |
| Listen, act and feedback | Good communication channels help address issues such as power, let people know what you are doing with their suggestions and why, ensure accountability. |
| Sharing information, experiences and knowledge | Sharing should be common across all groups involved. |
| Clarifying and agreeing expectations upfront | Clarify expectations to avoid conflicts, demotivation, dissolution of partnerships, or frustration in situations where stakeholders could perceive a lack of concrete actions, patients are 'partners' not 'are involved'. |
| Prioritising personal experience | Ensure people are clear on their role. |
| Scepticism | Avoid creating an atmosphere of doubt. |
| **Impact** | |
| **Heading** | **Aspects to consider** |
| Better evaluation | Develop and use evaluation throughout the project which focuses on PPI activity. |
| Continuous assessment and feedback | Impact should be measured across all groups involved. |
| Impact of project, PPI role and translation to real world situations | Monitor and reflect on impact throughout project. |
